# Evaluating the Efficacy of AI-Based Interactive Assessments Using Large Language Models for Depression Screening

**DOI:** 10.1101/2024.07.19.24310543

**Authors:** Zheng Jin, Dandan Bi, Jiaxing Hu, Kaibin Zhao

## Abstract

The evolution of language models, particularly the development of Large Language Models like ChatGPT, has opened new avenues for psychological assessment, potentially revolutionizing the rating scale methods that have been used for over a century. This study introduces a new Automated Assessment Paradigm (AAP), which aims to integrate natural language processing (NLP) techniques with traditional measurement methods. This integration enhances the accuracy and depth of mental health evaluations, while also addressing the acceptance and subjective experience of participants—areas that have not been extensively measured before. A pilot study was conducted with 32 participants, seven of whom were diagnosed with depression by licensed psychiatrists using the Clinical Interview Schedule-Revised (CIS-R). The participants completed the BDI-Fast Screen (BDI-FS) using a custom ChatGPT (GPTs) interface and the Chinese version of the PHQ-9 in a private setting. Following these assessments, participants also completed the Subjective Evaluation Scale. Spearman’s correlation analysis showed a high correlation between the total scores of the PHQ-9 and the BSI-FS-GPTs. The agreement of diagnoses between the two measures, as measured by Cohen’s kappa, was also significant. BSI-FS-GPTs diagnosis showed significantly higher agreement with the current diagnosis of depression. However, given the limited sample size of the pilot study, the AUC value of 1.00 and a sensitivity of 0.80 at a cutoff of 0.5, with zero false positive rate, likely overstate the classifier’s performance. Bayesian factors suggest that participants may feel more comfortable expressing their true feelings and opinions through this method. For ongoing follow-up research, a total sample size of approximately 104 participants, including about 26 diagnosed individuals, may be required to ensure the analysis maintains a necessary power of 0.80 and an alpha level of 0.05. Nonetheless, these findings provide a promising foundation for the ongoing validation of the new AAP in larger-scale studies, aiming to confirm its validity and reliability.

## INTRODUCTION

Numerous studies have employed numerical rating scales to capture participants’ feelings using predefined scoring responses to represent complex psychological states. These rating scales have yielded significant findings in psychology and other fields. When asked certain questions (e.g., “Are you satisfied with your partner?”), people often provide descriptive, open-ended responses in words (e.g., “Most of my expectations are met, but…”), rather than closed numerical or categorical answers (e.g., “7=*strongly agree*” or “1=*strongly disagree*”). The Likert (1932) scale format attempts to capture a unidimensional latent variable of individual attitudes by reducing potential responses to a relatively narrow, fixed range with limited resolution.

After years of development, advancements in classical test theory and item response theory have introduced better methods for selecting questions and aggregating closed-ended responses into latent variables that better represent actual states and characteristics (Heinen, 1996). However, the closed nature prevents respondents from flexibly expressing psychological states that do not fully align with the questions, leading to the neglect of complex or unusual perspectives. It is challenging to determine how respondents interpret the questions, what aspects they base their ratings on, the underlying cognitive processes, or whether they consider factors like family background, economic situation, or interpersonal relationships. Therefore, these methods are still limited by the inherent information loss in item response formats (Kjell, & Schwartz, 2024). Compared to open-ended natural language, rating scale methods appear overly simplified, despite being in use for nearly a century.

Natural language is our inherent way of conveying inner experiences and psychological states, characterized by high ecological validity, richness of information, and advantageous measurement properties (e.g., resolution, multidimensionality, and openness) (Lyons, 1991; Kjell, Kjell, & Schwartz, 2024). Language models have evolved from statistical (e.g., N-gram) to neural (e.g., Transformer). A few years ago, natural language systems were primarily tailored for specific tasks, such as sentiment analysis or generating alternative phrases for small portions of text, making the accurate quantification of language a challenging task (e.g., Lynn et al., 2018). In recent years, AI-based language analysis has fundamentally revolutionized the development of systems in the field. Now, with the rapid development and application of large language models like ChatGPT, using “instruction fine-tuning + reinforcement learning from human feedback” to align model outputs with human preferences has become foundational in natural language processing (Ke et al., 2023), enabling long-text analysis and modeling as well as multimodal data processing.

Natural language processing and machine learning address the challenge of translating language into scales. Recent studies by Kjell and colleagues have explored the quantitative assessment of language using AI-based models, validating the effectiveness of natural language processing techniques in mental health assessment. They used Latent Semantic Analysis (LSA) and context-based word embedding techniques (such as BERT) to analyze freely generated words and text responses. Findings showed that these techniques are highly consistent with traditional psychometric measurements (e.g., rating scales) (Kjell et al., 2019; 2021; Kjell 2022). Son et al. (2021) used AI language analysis techniques to predict the development of PTSD symptoms in 911 responders, highlighting early interview language features, including relative frequencies of words and phrases, binary indicators, and topic prevalence scores generated through Latent Dirichlet Allocation (LDA).

Natural language-based assessments show great potential in surpassing traditional rating scales, promising more accurate and comprehensive mental health evaluations. However, this technology brings not only ethical considerations (Bommasani et al., 2021) but also several other limitations. Most studies employed interviews to solicit open-ended responses or required participants to provide open-ended responses to a single question, transcribing and comparing the results of natural language analysis or classification to existing scale measurements for validation (e.g., Justyna Sarzynska-Wawer, 2021; Oltmanns et al., 2021) (Figure 1a). These studies validate the effectiveness of various models but do not explore how NLP techniques can be integrated with existing measurement methods to create more dynamic and adaptive assessment paradigms. AI programs, through complex prompts, facilitate the integration of delivering questions, collecting linguistic information, analyzing semantics, and scoring natural language (Figure 1b), presenting new opportunities for evaluating the new automated assessment paradigm (AAP). Furthermore, the acceptance and subjective experience of subjects using this technology remain unexplored, which is crucial for determining whether large language models for psychological assessment can be widely applied in the future.

**Figure 1.**
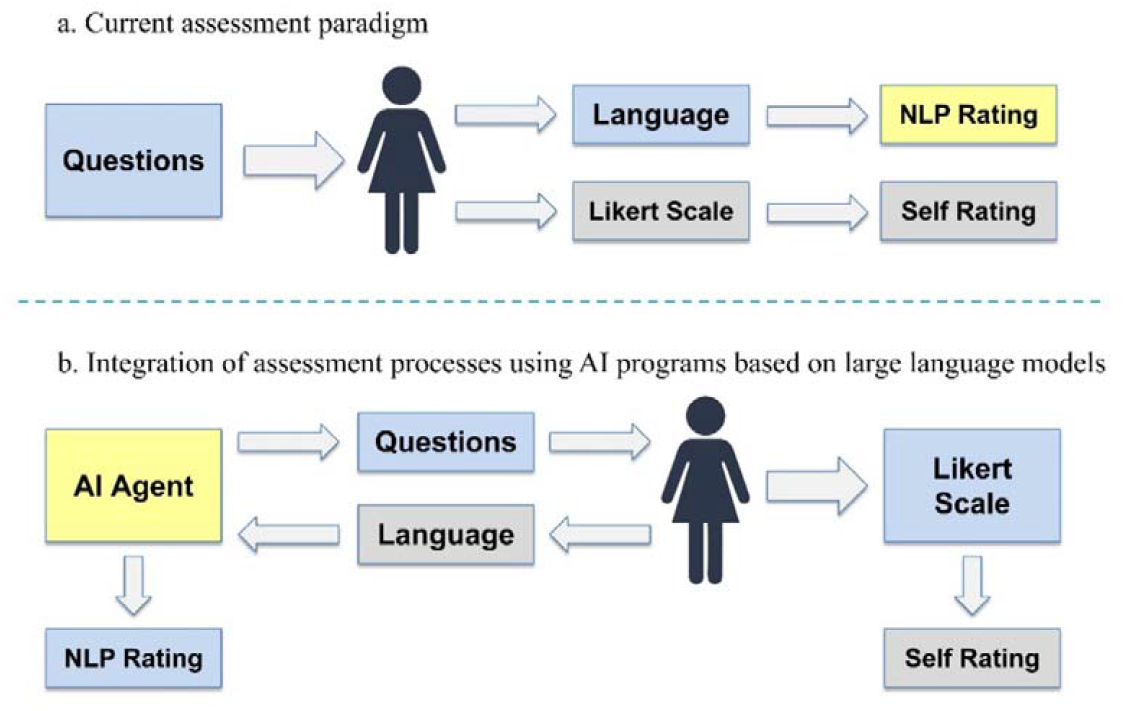
Current Effective Assessment Paradigm and Proposed Integration Paradigm.

## METHOD (Pilot study)

### Participants (Pilot study)

From the International Joint Laboratory of Psychological Science and the psychiatric outpatient department of a medical institution in Zhengzhou, 32 participants were recruited consecutively. Patients were Chinese native speaking, at least 18 years old. Seven participants were classified into the depression group, diagnosed by two licensed psychiatrists according to the “Diagnosis and Treatment of Mental Disorders guidelines (2020 Edition) “ issued by the National Health Commission of the People’s Republic of China. The diagnosis was based on the Clinical Interview Schedule-Revised (CIS-R) and supplemented by the 24-item Hamilton Depression Rating Scale (HAMD) (≥20). Current depressive episodes were diagnosed using an algorithm applying the ICD-10 classification system from CIS-R scores. Three participants were excluded due to cognitive or behavioral impediments that interfered with interviews and psychological testing, such as an inability to interact with GPTs using a keyboard. Five participants withdrew their participation in the study. Data from four participants were excluded from analysis due to unsuccessful completion of the program (e.g., sudden bugs or network interruptions). None of the participants had serious physical illnesses.

**Table 1.** *Demographic Data of Participants*.

|  | **Total (N = 20)** |
| --- | --- |
| **Gender, N(%)** |  |
| Male | 17 (85%) |
| Female | 3 (15%) |
| **Mean Age (SD)** | 23.40 (6.84) |
| **Age Range** | 18-41 |
| **Urban Area (N(%))** | 16 (80%) |
| **Rural Area (N(%))** | 4 (20%) |

*Note: Urban Area refers to household registration in urban regions, and Rural Area refers to household registration in rural regions.*

### Material

#### Beck Depression Inventory-II Fast Screen

(BDI-FS, Beck et al., 2000) is an extract from the 21-item BDI-II with only seven items. It has been validated for use with the general population, college students, and inpatients (e.g., Sören Kliem 2014; Elben et al., 2021), as well as its application within the Chinese population (e.g., Wong, 2011). Diagnostic cut-off value for BDI-FS is ≥4. Based on the BDI-SF, we created custom versions of ChatGPT (GPTs) on June 7, 2024 (for more information, https://openai.com/index/introducing-gpts/). Specifically, we used the original questions from the BDI-SF as a framework, prompting the GPTs to ask each question sequentially, waiting for the respondent to fully answer before moving on to the next.

The GPTs provided only courteous replies without commenting on the answers. For example, regarding self-criticalness in the BDI-SF, the prompt requires GPTs, acting as professional psychological assessors, to ask the respondent three questions sequentially: “Compared to before, do you find yourself more self-critical?”, “Do you blame yourself for your shortcomings?”, “When something bad happens, do you worry it’s your fault?” The GPTs then aggregate the respondent’s answers to these three questions for semantic recognition and analysis, following the scoring criteria of the original scale: I do not blame myself (0 points), I blame myself more than before (1 point), I blame myself for my faults (2 points), I blame myself for anything bad that happens (3 points) (Figure 2_right). We adapted the entire BDI-SF into an open-ended ten-question dialogue format, making them ideal for AI systems to capture nuanced linguistic cues (the tenth question is unrelated to the measurement). Two psychiatrists reviewed these questions. The GPTs automatically generate corresponding scores for seven of the original scale’s items based on the answers to nine of the questions. A complete example of the program is shown in Figure 2_left. Due to the information in Figure 2 being in a non-English language, it can be found at the following URLs: https://chatgpt.com/share/358b7517-7872-483d-919b-5236a9e9946e or https://osf.io/atq53/

### Patient Health Questionnaire-9 (PHQ-9)

The PHQ, developed by Kroenke et al. (2001) based on DSM-IV, is a self-assessment tool for evaluating mental disorders in primary healthcare settings. The PHQ-9 is a subset of the PHQ used specifically to assess depression, consisting of nine items based on the nine symptom criteria for major depressive disorder as outlined in the DSM-IV. Regarding severity, PHQ-9 comprises five categories, where a cut-off point of 0–4 indicates no depressive symptoms, 5–9 mild depressive symptoms, 10–14 moderate depressive symptoms, 15–19 moderately-severe depressive symptoms, and 20–27 severe depressive symptoms.

### Subjective Evaluation Scale

The subjective evaluation, revised from the Subjective Usability Scale in the dimensions of Usefulness, Ease of Use, Ease of Learning, and Satisfaction (e.g., Gao, Kortum, & Oswald, 2018), includes five items. These measure satisfaction with the information obtained, the degree to which the process felt considerate, and the extent to which the measurement method made the respondent feel respected. The scale includes three reverse-scored items, with higher scores indicating greater comfort.

### Procedure

All participants joined the study after signing informed consent forms and were free to withdraw at any time with no obligation to provide a reason. Demographic variables recorded included gender, age, and residence. All participants completed the BSI-FS-GPTs and the Chinese version of the PHQ-9 in a private space, guided by research assistants who were unaware of the diagnostic results. The order of the two assessments was counterbalanced, with a five-minute interval between them. The assessment end when the GPTs displayed the following concluding statements and final question, “Our conversation ends here. Thank you for your participation. Would you like to see your scores?” participants exited the program. The research assistant then reviewed and recorded the scores. After completing the two assessments, participants took an 8 to 10-minute break before completing the Subjective Usability Scale. Two research assistants were involved in the study.

### Result

Cronbach’s alpha for the PHQ-9 was 0.73, and for the BSI-FS-GPTs, it was 0.71. Spearman’s correlation analysis showed a high correlation between the total scores of the PHQ-9 and the BSI-FS-GPTs, with a correlation coefficient of 0.93. The agreement of diagnoses between the two measures, as measured by Cohen’s kappa, was significant, with a value of 0.83 (75%, *p* < 0.001). The BDI-SF diagnosis showed significantly higher agreement with the current diagnosis of depression, with a Cohen’s kappa of 0.86 (80%, *p* < 0.001). The PHQ-9 diagnosis also agreed significantly more often than chance with a present diagnosis of depression, as indicated by a significant Cohen’s kappa of 0.69 (60%, p < 0.001). The AUC value was 1.00, and at a cutoff of 0.5, the sensitivity was 0.80, with zero false positive rate, indicating that 80% of actual depression cases were correctly identified without any non-depressed cases being misclassified as depressed. However, considering the relatively small sample size, the practical significance of these results is limited (see Discussion).

Despite having only 20 participants, the results of the Bayesian T-test provided evidence supporting a difference in satisfaction between the two assessment methods, although the evidence was weak (Hu et al., 2018). *BF*_10_ = 1.27. There is a possibility that individuals may be more satisfied with the automated assessment paradigm (AAP) powered by GPT-based language models compared to traditional scoring assessments.

## DISCUSSION

Both measurements demonstrated acceptable internal consistency. There was a very high correlation between the PHQ-9 and the BSI-FS-GPTs, indicating a high level of agreement in measuring depressive symptoms. This result enhances confidence in the consistency and validity of both measurements under the same construct. The diagnostic results of both measurements showed significantly higher agreement with actual diagnoses than random chance. The agreement between the BDI-SF and actual diagnoses was very high. Given the limited sample size in the pilot study, with only 5 confirmed cases, statistical results are likely subject to significant fluctuations. The AUC value of 1.00 likely overstates the classifier’s actual performance. In a larger sample, a decrease in the AUC value would reflect a more realistic classification.

Based on previous studies using the BDI-SF in a sample of college students (Alsaleh & Lebreuilly, 2017), and considering the prevalence of diagnosed depression among individuals seeking psychological assistance (e.g., Kohn et al., 2004) and psychiatric outpatients (e.g., Yang et al., 2015), we set the ratio of sample sizes in the negative/positive groups to 3 in this study, with an expected acceptable value for the AUC (Area Under the Receiver Operating Characteristic Curve) of 0.80. A total sample size of approximately 104 participants, including about 26 diagnosed individuals, is required for Follow-Up Research, ensuring the analysis maintains the necessary power of 0.80 and an alpha level of 0.05.

The assessment based on virtual avatars reduces the need for human intervention, enhancing the efficiency of auxiliary diagnoses in medical institutions. With the future integration of AI technologies, questions adapted from classic scales may be presented in multimodal forms. As the processing and analysis of multimodal data continue to mature, this approach becomes increasingly viable. Participants are likely to feel more comfortable expressing their true feelings and opinions in this manner, and our pilot study data seem to support this expectation.

Overall, The current findings provide a promising foundation for the ongoing validation of the validity and reliability of the new Automated Assessment Paradigm (AAP) in larger-scale studies.

## Data Availability

All data produced are available online at https://osf.io/atq53/

